# Remote Ischemic Perconditioning and 90-Day Cognitive Outcomes After Acute Ischemic Stroke: A REMOTE-CAT Substudy

**DOI:** 10.64898/2026.07.08.26357601

**Authors:** Cristina Pereira-Priego, Gloria Arqué, Ares Regué, Gerard Mauri-Capdevila, Xavier Jiménez-Fàbrega, Teresa Subirats, José Ramón Ropero, Mikel Vicente-Pascual, Àlex Rovira, Sara Salvany, Cristina García-Vázquez, Rafel Cirer-Sastre, Francisco Purroy, the REMOTE-CAT Trial Investigators

## Abstract

**Background:** Remote ischemic perconditioning (RIperC) is a simple, noninvasive neuroprotective strategy based on brief cycles of limb ischemia-reperfusion during cerebral ischemia. REMOTE-CAT suggested a potential functional benefit of prehospital RIperC in acute ischemic stroke. However, its effect on poststroke cognitive outcomes, which may not be fully captured by global disability scales, remains uncertain.

**Methods:** We performed an exploratory cognitive substudy of the multicenter, randomized, double-blind, sham-controlled REMOTE-CAT trial. Patients with suspected acute ischemic stroke within 8 hours, prestroke modified Rankin Scale score <3, and RACE motor score >0 were randomized prehospital to RIperC or sham. RIperC consisted of five 5-minute cuff inflation-deflation cycles during ambulance transfer. At 90 days, patients from one center underwent a standardized neuropsychological battery assessing five cognitive domains.

**Results:** Among 122 patients in the primary analysis, 58 (47.5%) completed neuropsychological assessment: 26 allocated to RIperC and 32 to sham. No statistically significant between-group differences were observed in domain-specific Z scores. Cognitive impairment in at least one domain was numerically less frequent with RIperC than sham (26.9% versus 34.4%). Impairment in more than one domain was also less frequent with RIperC (7.7% versus 21.9%), although the overall distribution of impaired domains did not differ significantly between groups (P=0.244). The largest domain-specific difference was observed for visual memory impairment (3.8% versus 21.9%).

**Conclusions:** In this exploratory substudy, prehospital RIperC did not significantly improve 90-day cognitive outcomes after acute ischemic stroke. Nevertheless, RIperC-treated patients showed numerically favorable trends, particularly in global cognitive burden and visual memory. These hypothesis-generating findings support incorporating standardized cognitive outcomes in future ischemic conditioning trials.

**Clinical Trial Registration:** NCT03375762.

## Introduction

Cognitive impairment is a frequent and clinically meaningful consequence of stroke. It may affect independence, return to work, driving, social participation, interpersonal relationships, and quality of life. Importantly, poststroke cognitive impairment may occur even among patients with mild stroke or apparently favorable functional recovery.^1,2^ Therefore, cognitive outcomes provide complementary information to conventional global disability scales and may be particularly relevant when evaluating neuroprotective interventions.

The modified Rankin Scale (mRS) remains the standard outcome measure in acute stroke trials and is widely used as a global disability endpoint.^3^ However, the mRS is relatively insensitive to deficits in memory, attention, processing speed, executive function, visuospatial abilities, language, and praxis. A patient may be classified as functionally independent according to the mRS while still experiencing cognitive deficits that substantially affect daily functioning.^2^ Accordingly, cognitive outcomes are increasingly being incorporated into acute stroke trials and secondary analyses of reperfusion therapies. Cognitive substudies of endovascular thrombectomy trials, including REVASCAT and ESCAPE, have shown that acute reperfusion treatment may be associated with better cognitive performance beyond its effect on global disability scales.^4,5^ Similarly, analyses from DEFUSE 3 showed improvements in quality-of-life domains including cognitive function after endovascular therapy.^6^ These studies support the systematic assessment of cognitive outcomes as complementary endpoints in acute stroke trials.

Remote ischemic conditioning (RIC) is a noninvasive intervention based on brief, controlled episodes of limb ischemia and reperfusion aimed at activating endogenous protective pathways in distant organs.^7,8^ When applied during ongoing cerebral ischemia, it is referred to as remote ischemic perconditioning (RIperC).^8^ Experimental data suggest that ischemic conditioning may modulate several mechanisms involved in ischemic brain injury and recovery, including inflammation, oxidative stress, endothelial dysfunction, mitochondrial injury, apoptosis, blood-brain barrier disruption, microcirculatory impairment, and neurorepair pathways.^7,8^ These mechanisms provide a biological rationale for evaluating whether RIperC may influence not only infarct evolution and global functional outcome, but also poststroke cognition.^9,10^

Clinical evidence on RIC in acute ischemic stroke remains heterogeneous. A recent systematic review and meta-analysis of randomized trials suggested that the effects of RIC may differ according to the concomitant acute treatment strategy, with more favorable signals in medically treated patients and less consistent benefit among patients receiving intravenous thrombolysis or mechanical thrombectomy.^11^ These findings highlight the importance of treatment timing, protocol intensity, population selection, and outcome selection in ischemic conditioning trials. In this context, the REMOTE-CAT trial evaluated an early prehospital RIperC strategy in patients with suspected acute ischemic stroke and suggested a potential functional benefit, supporting further exploration of outcomes beyond global disability.^12^

Whether prehospital RIperC improves cognitive outcomes after acute ischemic stroke remains unknown. We conducted an exploratory cognitive substudy of the REMOTE-CAT randomized clinical trial to evaluate whether prehospital RIperC was associated with better 90-day cognitive performance across prespecified cognitive domains.

## Methods

### Study Design

This was a cognitive substudy of the REMOTE-CAT trial, an investigator-initiated, multicenter, randomized, double-blind, sham-controlled clinical trial conducted in Catalonia, Spain.^12,13^ REMOTE-CAT evaluated the effect of prehospital RIperC in patients with suspected acute ischemic stroke.^12,13^ The parent trial was registered at https://ClinicalTrials.gov (NCT03375762). The present substudy included patients recruited at Hospital Universitari Arnau de Vilanova de Lleida, Spain, one of the four participating centers in the REMOTE-CAT trial, where a standardized neuropsychological assessment was performed at 90 days after the index event. This analysis was designed as a hypothesis-generating substudy.

### Ethics Approval and Consent to Participate

The REMOTE-CAT trial was approved by the Ethics Committee of Hospital Universitari Arnau de Vilanova de Lleida, Spain (code 1744), as central ethics committee, and by the ethics committee of each participating center. The trial also received approval from the Spanish Medical Agency and was conducted in accordance with the Declaration of Helsinki and Good Clinical Practice guidelines. The protocol followed CONSORT 2010 guidelines and was registered at https://ClinicalTrials.gov (NCT03375762). Written informed consent was obtained from all participants or their legal representatives, according to the procedures approved for the parent trial, including deferred consent when immediate consent was not possible in the prehospital or acute stroke setting.

### Participants

The parent trial enrolled adult patients evaluated in the prehospital setting for suspected acute ischemic stroke. Main inclusion criteria were age >18 years, symptom onset within 8 hours, prestroke mRS score <3, and motor symptoms at inclusion, defined as a RACE motor score >0, as previously described in the REMOTE-CAT trial protocol and primary publication.^12,13^ Patients with unknown symptom onset, coma, pregnancy, participation in another clinical trial, or malignant disease or severe comorbidity associated with life expectancy <6 months were excluded.^12,13^

For the present substudy, eligible patients were those included in the primary REMOTE-CAT analysis who completed the 90-day neuropsychological battery.

### Randomization and Masking

In the parent trial, patients were randomized in a 1:1 ratio to active RIperC or sham using a centralized allocation system. Randomization was stratified according to prehospital RACE score. Active and sham devices were externally identical and produced similar noise and vibration, preserving blinding of patients, clinical investigators, and outcome assessors. Neuropsychological assessment was performed by trained personnel blinded to treatment allocation.

### Intervention

RIperC was initiated in the ambulance immediately after randomization. In the active treatment group, an automated cuff was placed on the upper nonaffected limb and inflated to 200 mmHg for 5 minutes, followed by 5 minutes of deflation. The protocol consisted of five inflation-deflation cycles. If required, the procedure was continued after hospital arrival until completion. In the sham group, a sham cuff simulated the active device without producing therapeutic ischemia. All patients received standard acute stroke care according to local and international guidelines, including reperfusion therapies when indicated.

### Neuropsychological Assessment

At 90 days after stroke, patients underwent a standardized neuropsychological assessment administered by trained neuropsychology personnel blinded to treatment allocation. The battery evaluated five prespecified cognitive domains. Verbal memory was assessed using the Buschke Test.^14^ Visual memory was evaluated with the recall condition of the Rey-Osterrieth Complex Figure Test, whereas constructional praxis was assessed using its copy condition.^15^ Attention and information processing speed were assessed using Trail Making Test A and B,^16^ Forward Digit Span from the Spanish adaptation of the WAIS-IV,^17^ direct spatial localization from the Barcelona Test,^18^ and the copy time of the Rey-Osterrieth Complex Figure Test.^15^ Executive function was assessed using phonemic fluency with the letters P, M, and R, semantic fluency using the animal category, Backward Digit Span from the Spanish adaptation of the WAIS-IV,^17^ and indirect spatial localization from the Barcelona Test.^18^ The neuropsychological tests used to define each cognitive domain are summarized in Supplementary Table 2.

Raw neuropsychological test scores were first converted into scaled scores using NEURONORMA normative data, adjusted for age and educational level when such corrections were available.^19–22^ When education-adjusted norms were not available for a given measure, the corresponding age-adjusted normative score was used. Scaled scores were subsequently transformed into domain-specific Z scores, allowing standardized comparisons across cognitive domains. For each domain, cognitive impairment was defined according to prespecified normative thresholds. Cognitive impairment was analyzed both as domain-specific impairment and as global cognitive burden, defined by the number of impaired cognitive domains. The proportion of patients with impairment in at least one domain and in more than one domain was also assessed.

### Clinical and Neuroimaging Variables

Baseline demographic and clinical variables included age, sex, vascular risk factors, educational level, prestroke mRS, prehospital RACE score, admission NIHSS score, stroke etiology, and acute phase treatments. Neuroimaging variables included ASPECTS, presence and site of LVO, DWI lesion volume, vascular territory, Fazekas score, cerebral microbleeds, and brain atrophy.

Neuroimaging variables were assessed by an independent centralized imaging core laboratory blinded to treatment allocation and clinical data, following the procedures of the parent REMOTE-CAT trial. The standardized imaging protocol included noncontrast brain CT, CT angiography with source images and maximum intensity projection reconstructions in axial and coronal planes, T2-FLAIR, T2*-weighted imaging, 3D time-of-flight MR angiography, diffusion-weighted imaging with b=1000 s/mm², and ADC maps. The MRI protocol and infarct volume quantification methods are described in the Supplementary Methods. Brain MRI was performed within 7 days of symptom onset when available. DWI lesion volume was calculated using OLEA Sphere software, with expert verification and manual adjustment of segmented infarct areas when needed. White matter hyperintensity burden was graded using the Fazekas scale. Cerebral microbleeds were defined according to established small vessel disease neuroimaging standards as small, rounded or ovoid hypointense lesions on T2*-weighted imaging, compatible with hemosiderin deposits and distinct from vascular flow voids, calcification, or other mimics.^23^ Brain atrophy was visually rated using the Global Cortical Atrophy scale and categorized into three levels: no atrophy, mild atrophy, and moderate-to-severe atrophy.^24^

### Outcomes

The primary outcome of this substudy was 90-day cognitive performance across the five prespecified neuropsychological domains: verbal memory, visual memory, constructional praxis, attention and information processing speed, and executive function. Secondary outcomes included the proportion of patients with impairment in each cognitive domain, the number of impaired cognitive domains, impairment in at least one domain, and impairment in more than one domain.

Functional outcome in the substudy was assessed using the same definition as in the parent REMOTE-CAT trial: favorable functional outcome was defined as mRS score <3 at 90 days.

### Statistical Analysis

This substudy was not powered to detect statistically significant between-group differences in cognitive outcomes. The analysis population included patients from the REMOTE-CAT primary efficacy population who completed the 90-day neuropsychological assessment.

Baseline characteristics were summarized according to treatment allocation. Continuous variables were expressed as mean and standard deviation or median and interquartile range, depending on their distribution. Normality was assessed using the Shapiro-Wilk test. Categorical variables were summarized as counts and percentages. Between-group comparisons were performed using Student’s t test or the Mann-Whitney U test for continuous variables, and Pearson’s chi-square test or Fisher’s exact test for categorical variables, as appropriate. Neuropsychological performance at 90 days was analyzed using domain-specific Z scores. Between-group differences in Z scores were assessed using the Mann-Whitney U test because of the limited sample size. Cognitive impairment was analyzed as a binary outcome for each domain according to prespecified impairment thresholds. For binary cognitive outcomes, unadjusted odds ratios with 95% confidence intervals were estimated using logistic regression. To maintain consistency with the statistical approach of the parent REMOTE-CAT trial, covariate-adjusted analyses used a parsimonious model including age, prestroke mRS score, and baseline stroke severity assessed by the prehospital RACE score. Given the small number of events in several cognitive domains, adjusted estimates were interpreted cautiously. Functional outcome in the substudy was evaluated using logistic regression models. Favorable functional outcome was defined as mRS score <3 at 90 days, as in the parent REMOTE-CAT trial. The covariate-adjusted model included age, prestroke mRS score, and prehospital RACE score, consistent with the post hoc adjusted analysis of the primary REMOTE-CAT trial. Because all neuropsychological analyses were hypothesis-generating, no correction for multiplicity was applied across cognitive domains or secondary cognitive outcomes. Therefore, domain-specific P values should be interpreted descriptively rather than as confirmatory evidence. All statistical tests were two-sided, and a P value <0.05 was considered nominally significant. Statistical analyses were performed using SPSS version 27.0 and R version 4.1.0.

## Results

### Study Population

The flow of participants through the cognitive substudy is shown in Figure 1. Between August 2019 and December 2023, 350 patients were assessed for eligibility, 220 were enrolled, and 200 were randomized in the REMOTE-CAT trial. Among randomized patients, 93 received RIperC and 107 received sham treatment. After exclusion of patients with stroke mimics, intracerebral hemorrhage, or prestroke mRS score >2, 122 patients were included in the primary REMOTE-CAT analysis: 57 in the RIperC group and 65 in the sham group. Among patients included in the primary analysis, 51 patients in the RIperC group and 55 patients in the sham group completed the 90-day follow-up. The 90-day neuropsychological assessment was completed by 58 of 122 patients included in the primary REMOTE-CAT analysis (47.5%), who constituted the cognitive substudy population: 26 allocated to RIperC and 32 allocated to sham.

**Figure 1.**
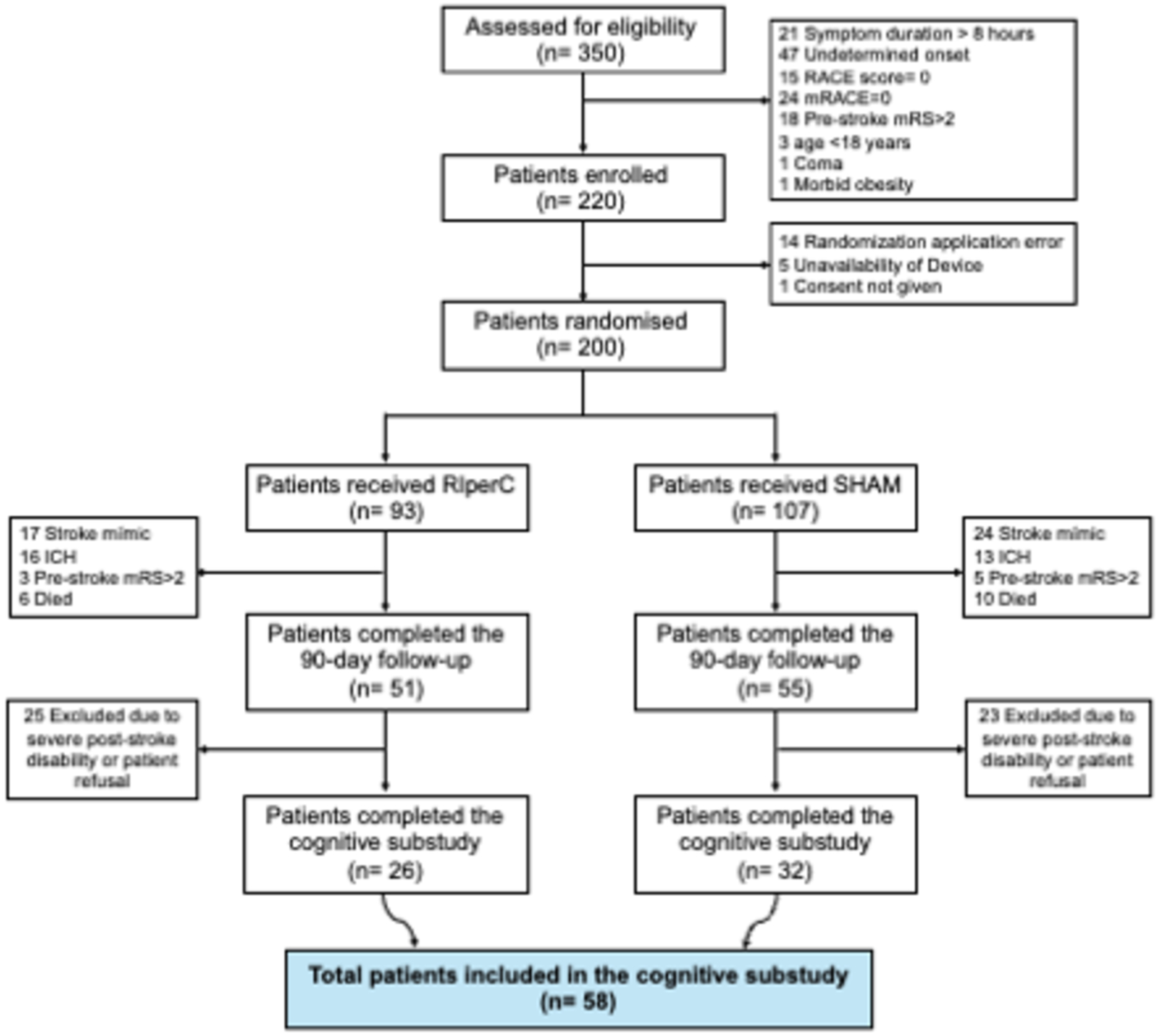
Flowchart of participants included in the cognitive substudy of the REMOTE-CAT trial. Flow chart showing eligibility assessment, enrollment, randomization, inclusion in the primary REMOTE-CAT analysis, 90-day follow-up, and inclusion in the cognitive substudy. ICH indicates intracerebral hemorrhage; mRS, modified Rankin Scale; RIperC, remote ischemic perconditioning.

Baseline characteristics, treatment allocation, and 90-day functional outcome of patients included and not included in the cognitive substudy are shown in Supplementary Table 1. Treatment allocation was well balanced between patients included and not included in the substudy: 26 of 58 included patients (44.8%) and 31 of 64 nonincluded patients (48.4%) had been allocated to RIperC, whereas 32 of 58 included patients (55.2%) and 33 of 64 nonincluded patients (51.6%) had been allocated to sham (P=0.690). Patients included in the substudy had lower admission NIHSS scores than those not included (median, 7.5 [IQR, 4-14] versus 10.5 [IQR, 4.5-20.5]; P=0.007) and a higher proportion of favorable functional outcome at 90 days (mRS <3: 67.2% versus 45.3%; P=0.015). No statistically significant differences were observed in age, sex, vascular risk factors, prestroke mRS score, prehospital RACE score, ASPECTS, LVO, stroke etiology, or acute reperfusion treatment.

### Baseline Characteristics

Baseline clinical and neuroimaging characteristics according to treatment allocation are shown in Table 1. The mean age of the substudy population was 70.0±15.0 years, and 22 patients were women (37.9%). Age, sex, prestroke mRS score, vascular risk factors, acute phase treatment, ASPECTS, presence of LVO, DWI lesion volume, Fazekas score, cerebral microbleed burden, and brain atrophy were broadly comparable between the RIperC and sham groups.

**Table 1.**
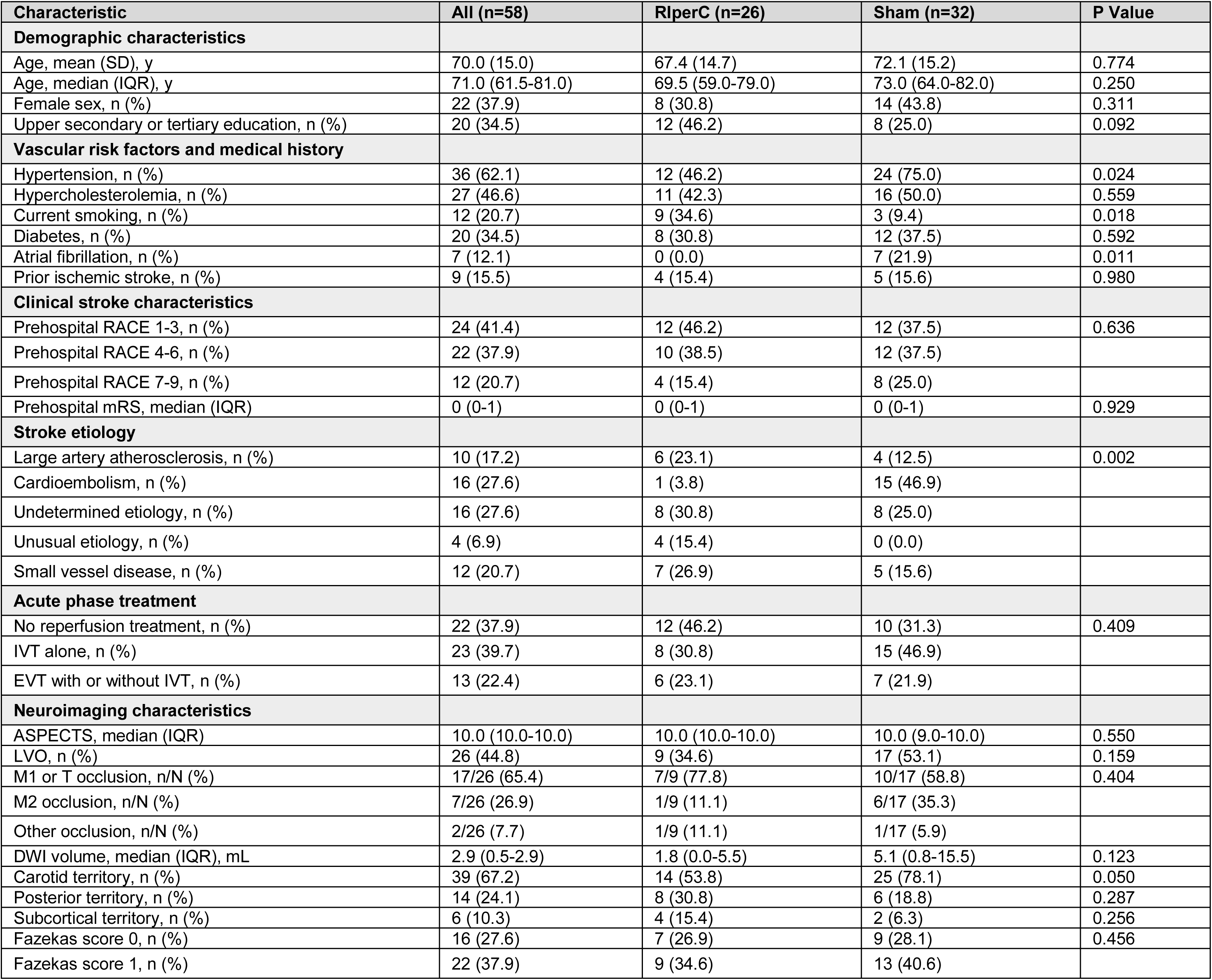
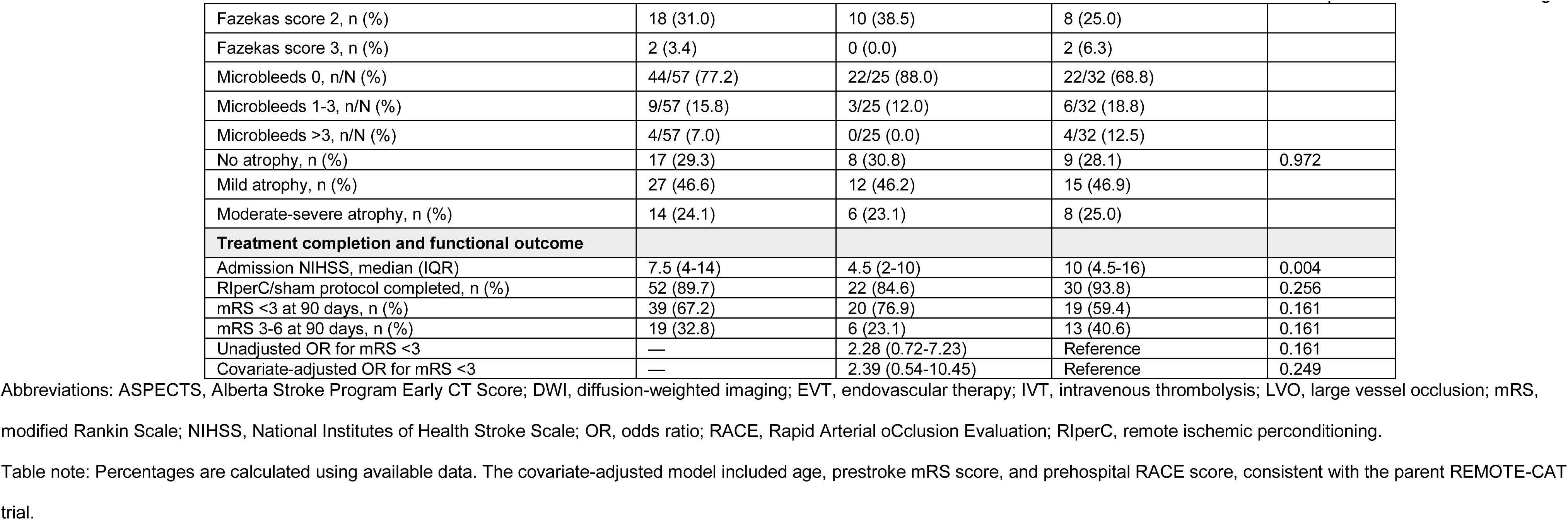
Baseline Clinical and Neuroimaging Characteristics of the Cognitive Substudy Population.

| Characteristic | All (n=58) | RlperC (n=26) | Sham (n=32) | P Value |
| --- | --- | --- | --- | --- |
| **Demographic characteristics** |  |  |  |  |
| Age, mean (SD), y | 70.0 (15.0) | 67.4 (14.7) | 72.1 (15.2) | 0.774 |
| Age, median (IQR), y | 71.0 (61.5-81.0) | 69.5 (59.0-79.0) | 73.0 (64.0-82.0) | 0.250 |
| Female sex, n (%) | 22 (37.9) | 8 (30.8) | 14 (43.8) | 0.311 |
| Upper secondary or tertiary education, n (%) | 20 (34.5) | 12 (46.2) | 8 (25.0) | 0.092 |
| **Vascular risk factors and medical history** |  |  |  |  |
| Hypertension, n (%) | 36 (62.1) | 12 (46.2) | 24 (75.0) | 0.024 |
| Hypercholesterolemia, n (%) | 27 (46.6) | 11 (42.3) | 16 (50.0) | 0.559 |
| Current smoking, n (%) | 12 (20.7) | 9 (34.6) | 3 (9.4) | 0.018 |
| Diabetes, n (%) | 20 (34.5) | 8 (30.8) | 12 (37.5) | 0.592 |
| Atrial fibrillation, n (%) | 7 (12.1) | 0 (0.0) | 7 (21.9) | 0.011 |
| Prior ischemic stroke, n (%) | 9 (15.5) | 4 (15.4) | 5 (15.6) | 0.980 |
| **Clinical stroke characteristics** |  |  |  |  |
| Prehospital RACE 1-3, n (%) | 24 (41.4) | 12 (46.2) | 12 (37.5) | 0.636 |
| Prehospital RACE 4-6, n (%) | 22 (37.9) | 10 (38.5) | 12 (37.5) |  |
| Prehospital RACE 7-9, n (%) | 12 (20.7) | 4 (15.4) | 8 (25.0) |  |
| Prehospital mRS, median (IQR) | 0 (0-1) | 0 (0-1) | 0 (0-1) | 0.929 |
| **Stroke etiology** |  |  |  |  |
| Large artery atherosclerosis, n (%) | 10 (17.2) | 6 (23.1) | 4 (12.5) | 0.002 |
| Cardioembolism, n (%) | 16 (27.6) | 1 (3.8) | 15 (46.9) |  |
| Undetermined etiology, n (%) | 16 (27.6) | 8 (30.8) | 8 (25.0) |  |
| Unusual etiology, n (%) | 4 (6.9) | 4 (15.4) | 0 (0.0) |  |
| Small vessel disease, n (%) | 12 (20.7) | 7 (26.9) | 5 (15.6) |  |
| **Acute phase treatment** |  |  |  |  |
| No reperfusion treatment, n (%) | 22 (37.9) | 12 (46.2) | 10 (31.3) | 0.409 |
| IVT alone, n (%) | 23 (39.7) | 8 (30.8) | 15 (46.9) |  |
| EVT with or without IVT, n (%) | 13 (22.4) | 6 (23.1) | 7 (21.9) |  |
| **Neuroimaging characteristics** |  |  |  |  |
| ASPECTS, median (IQR) | 10.0 (10.0-10.0) | 10.0 (10.0-10.0) | 10.0 (9.0-10.0) | 0.550 |
| LVO, n (%) | 26 (44.8) | 9 (34.6) | 17 (53.1) | 0.159 |
| M1 or T occlusion, n/N (%) | 17/26 (65.4) | 7/9 (77.8) | 10/17 (58.8) | 0.404 |
| M2 occlusion, n/N (%) | 7/26 (26.9) | 1/9 (11.1) | 6/17 (35.3) |  |
| Other occlusion, n/N (%) | 2/26 (7.7) | 1/9 (11.1) | 1/17 (5.9) |  |
| DWI volume, median (IQR), mL | 2.9 (0.5-2.9) | 1.8 (0.0-5.5) | 5.1 (0.8-15.5) | 0.123 |
| Carotid territory, n (%) | 39 (67.2) | 14 (53.8) | 25 (78.1) | 0.050 |
| Posterior territory, n (%) | 14 (24.1) | 8 (30.8) | 6 (18.8) | 0.287 |
| Subcortical territory, n (%) | 6 (10.3) | 4 (15.4) | 2 (6.3) | 0.256 |
| Fazekas score 0, n (%) | 16 (27.6) | 7 (26.9) | 9 (28.1) | 0.456 |
| Fazekas score 1, n (%) | 22 (37.9) | 9 (34.6) | 13 (40.6) |  |
| Fazekas score 2, n (%) | 18 (31.0) | 10 (38.5) | 8 (25.0) |  |
| Fazekas score 3, n (%) | 2 (3.4) | 0 (0.0) | 2 (6.3) |  |
| Microbleeds 0, n/N (%) | 44/57 (77.2) | 22/25 (88.0) | 22/32 (68.8) |  |
| Microbleeds 1-3, n/N (%) | 9/57 (15.8) | 3/25 (12.0) | 6/32 (18.8) |  |
| Microbleeds >3, n/N (%) | 4/57 (7.0) | 0/25 (0.0) | 4/32 (12.5) |  |
| No atrophy, n (%) | 17 (29.3) | 8 (30.8) | 9 (28.1) | 0.972 |
| Mild atrophy, n (%) | 27 (46.6) | 12 (46.2) | 15 (46.9) |  |
| Moderate-severe atrophy, n (%) | 14 (24.1) | 6 (23.1) | 8 (25.0) |  |
| **Treatment completion and functional outcome** |  |  |  |  |
| Admission NIHSS, median (IQR) | 7.5 (4-14) | 4.5 (2-10) | 10 (4.5-16) | 0.004 |
| RIperC/sham protocol completed, n (%) | 52 (89.7) | 22 (84.6) | 30 (93.8) | 0.256 |
| mRS <3 at 90 days, n (%) | 39 (67.2) | 20 (76.9) | 19 (59.4) | 0.161 |
| mRS 3-6 at 90 days, n (%) | 19 (32.8) | 6 (23.1) | 13 (40.6) | 0.161 |
| Unadjusted OR for mRS <3 | — | 2.28 (0.72-7.23) | Reference | 0.161 |
| Covariate-adjusted OR for mRS <3 | — | 2.39 (0.54-10.45) | Reference | 0.249 |

Abbreviations: ASPECTS, Alberta Stroke Program Early CT Score; DWI, diffusion-weighted imaging; EVT, endovascular therapy; IVT, intravenous thrombolysis; LVO, large vessel occlusion; mRS, modified Rankin Scale; NIHSS, National Institutes of Health Stroke Scale; OR, odds ratio; RACE, Rapid Arterial occlusion Evaluation; RIperC, remote ischemic preconditioning.
Table note: Percentages are calculated using available data. The covariate-adjusted model included age, prestroke mRS score, and prehospital RACE score, consistent with the parent REMOTE-CAT trial.

Importantly, prehospital stroke severity, assessed using the RACE score before treatment initiation, was similarly distributed between groups. Some baseline imbalances were observed: hypertension and atrial fibrillation were more frequent in the sham group, whereas current smoking was more frequent in the RIperC group. Stroke etiology also differed between groups, with cardioembolism more frequent in the sham group.

### Treatment Completion and Functional Outcome

Treatment completion, admission NIHSS score, and 90-day functional outcomes are shown in Table 1. The RIperC or sham protocol was completed in 22 patients in the RIperC group and 30 patients in the sham group (84.6% versus 93.8%; P=0.256).

Admission NIHSS scores were lower in the RIperC group than in the sham group (median, 4.5 [IQR, 2-10] versus 10 [IQR, 4.5-16]; P=0.004). A favorable functional outcome, defined as mRS <3 at 90 days, was observed in 20 patients in the RIperC group and 19 patients in the sham group (76.9% versus 59.4%; P=0.161). The unadjusted odds ratio for mRS <3 was 2.28 (95% CI, 0.72-7.23), and the covariate-adjusted odds ratio was 2.39 (95% CI, 0.54-10.45; P=0.249).

### Cognitive Impairment

Domain-specific cognitive performance, global cognitive burden, and domain-specific cognitive impairment at 90 days are shown in Figures 2 and 3. No statistically significant between-group differences were observed in Z scores across any of the five prespecified cognitive domains: verbal memory, visual memory, constructional praxis, attention and information processing speed, and executive function.

**Figure 2.**
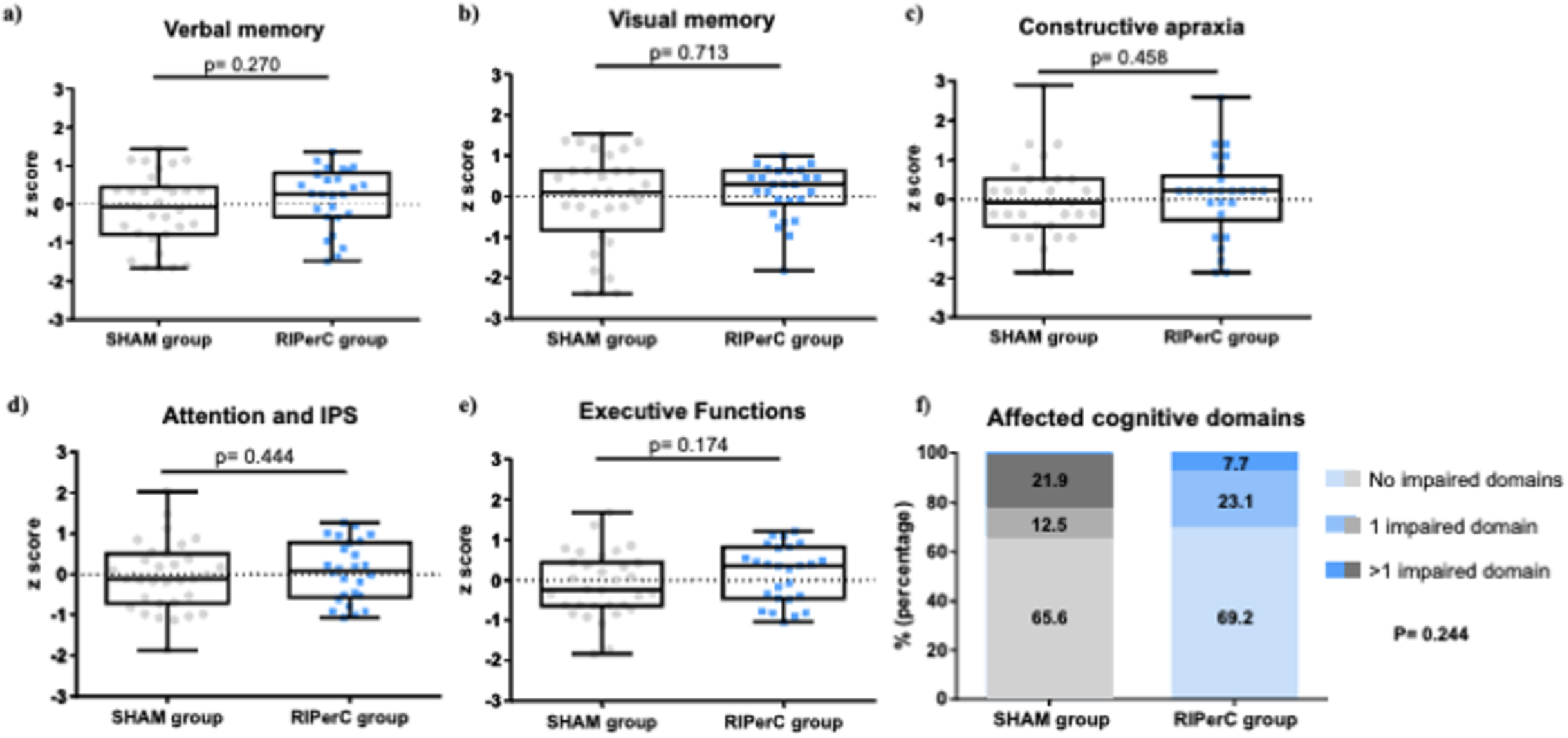
Domain-Specific Cognitive Outcomes and Global Cognitive Burden at 90 Days. Box-and-whisker plots showing domain-specific cognitive performance at 90 days, expressed as Z scores, in patients randomized to remote ischemic perconditioning (RIperC; n=26) or sham treatment (n=32). Higher Z scores indicate better cognitive performance. Panels A-E show verbal memory, visual memory, constructional praxis, attention and information processing speed, and executive function. Panel F shows global cognitive burden according to the number of impaired cognitive domains. Boxes represent the interquartile range, the central line indicates the median, and whiskers represent the range. Individual patient values are overlaid. Between-group comparisons for domain-specific Z scores were performed using the Mann-Whitney U test. RIperC indicates remote ischemic perconditioning.

**Figure 3.**
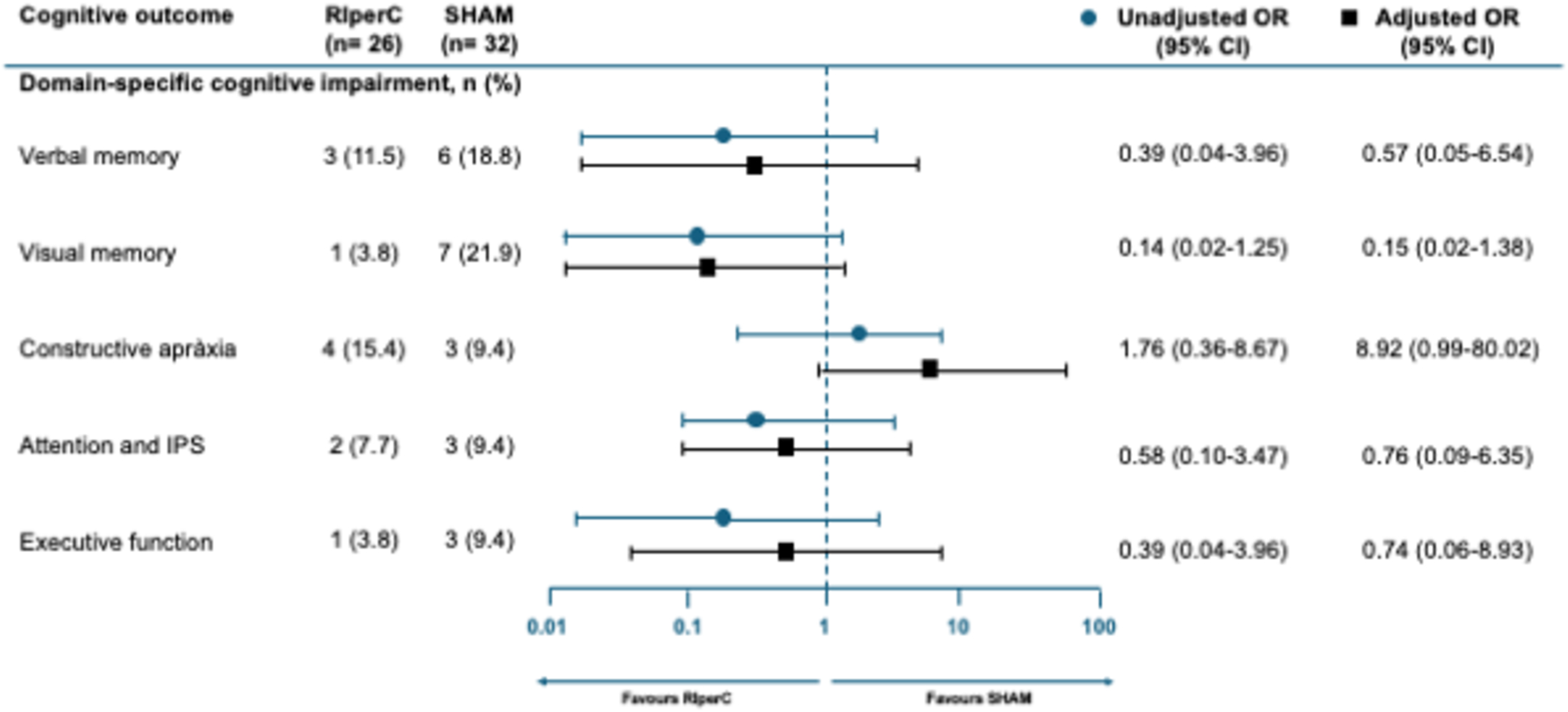
Effect of Remote Ischemic Perconditioning on Domain-Specific Cognitive Impairment at 90 Days. Forest plot showing the association between treatment allocation and domain-specific cognitive impairment at 90 days. For each cognitive domain, the number and percentage of patients with impairment are shown according to treatment group, together with unadjusted and covariate-adjusted odds ratios and 95% confidence intervals. Odds ratios compare RIperC with sham. The adjusted model included age, prestroke modified Rankin Scale score, and prehospital RACE score. IPS indicates information processing speed; OR, odds ratio; RIperC, remote ischemic perconditioning.

When cognitive outcomes were analyzed as domain-specific impairment, the proportion of patients with impairment was numerically lower in the RIperC group for verbal memory, visual memory, attention and information processing speed, and executive function. The largest absolute between-group difference was observed for visual memory impairment, which was present in 1 of 26 patients in the RIperC group and 7 of 32 patients in the sham group (3.8% versus 21.9%; unadjusted odds ratio, 0.14 [95% CI, 0.02-1.25]; adjusted odds ratio, 0.15 [95% CI, 0.02-1.38]). None of the domain-specific comparisons reached statistical significance. Constructional praxis impairment was observed in 4 of 26 RIperC-treated patients and 3 of 32 sham-treated patients (15.4% versus 9.4%). The adjusted estimate for constructional praxis impairment was unstable because of the small number of events and wide confidence intervals and should not be interpreted as evidence of harm.

When global cognitive burden was analyzed according to the number of impaired domains, impairment in more than one cognitive domain was observed in 2 of 26 RIperC-treated patients and 7 of 32 sham-treated patients (7.7% versus 21.9%). The overall distribution of impaired cognitive domains did not differ significantly between groups (P=0.244).

## Discussion

In this cognitive substudy of the REMOTE-CAT randomized clinical trial, prehospital RIperC was not associated with statistically significant differences in 90-day cognitive performance across prespecified cognitive domains. However, several numerical trends favored RIperC. Patients allocated to RIperC showed a lower proportion of impairment in most cognitive domains, with the largest absolute difference observed for visual memory impairment. In addition, impairment in more than one cognitive domain was less frequent in the RIperC group than in the sham group. These findings should be interpreted as hypothesis-generating given the modest sample size, the exploratory nature of the analyses, and the limited number of events in several cognitive outcomes.

Our results should be interpreted in the context of the growing recognition that cognitive outcomes provide information that is complementary to global disability scales in acute stroke trials. Prior cognitive substudies of reperfusion trials have shown that treatment effects may extend beyond conventional mRS-based outcomes and may be detectable in cognitive or patient-centered domains.^4–6^ In this setting, the present substudy extends the assessment of cognition to a prehospital neuroprotection strategy. Although RIperC was not associated with statistically significant cognitive benefit, the observed numerical differences support the relevance and feasibility of incorporating standardized cognitive outcomes in future ischemic conditioning trials.

The biological rationale for assessing cognitive outcomes after RIperC is supported by experimental and clinical evidence suggesting that ischemic conditioning may modulate pathways involved in ischemic injury and recovery, including inflammation, oxidative stress, endothelial dysfunction, mitochondrial injury, apoptosis, blood-brain barrier disruption, microcirculatory impairment, and neurorepair.^7,8^ These pathways may influence not only infarct evolution and global disability, but also cognitive recovery. Previous clinical evidence on RIC and cognition remains limited. A pilot randomized trial suggested that RIC may improve disability and cognition after acute ischemic stroke, and a systematic review of RIC and cognitive performance concluded that repeated protocols may have potential cognitive benefits in selected populations, whereas evidence from single-session interventions remains less consistent.^9,10^ In this context, the numerical differences observed in our study, particularly for visual memory and global cognitive burden, provide a signal that warrants further evaluation in adequately powered trials.

The clinical evidence for RIC in acute ischemic stroke remains heterogeneous. RESCUE BRAIN found no significant effect of in-hospital RIperC on infarct growth, whereas RESIST found no significant improvement in 90-day functional outcome after prehospital RIC continued during hospitalization.^25,26^ In contrast, RICAMIS reported improved 90-day neurological outcome with repeated RIC in patients with acute moderate ischemic stroke.^27^ The primary REMOTE-CAT trial suggested a favorable direction of effect for prehospital RIperC and showed a significant association with favorable functional outcome in a covariate-adjusted post hoc analysis.^12^ A recent systematic review and meta-analysis suggested that the effects of RIC may differ according to concomitant acute treatment strategy, with more favorable signals in medically treated patients and less consistent benefit among patients receiving intravenous thrombolysis or mechanical thrombectomy.^11^ Together, these data suggest that the effects of ischemic conditioning may depend on treatment timing, protocol intensity, patient selection, concomitant reperfusion therapy, and outcome definition.

The largest absolute difference in our substudy was observed for visual memory impairment. Although this finding did not reach statistical significance, visual memory may be particularly sensitive to distributed cortical-subcortical network injury, lesion location, and subtle poststroke cognitive dysfunction not captured by global disability scales. Nevertheless, this domain-specific signal should be interpreted cautiously. The substudy was not powered for individual cognitive domains, confidence intervals were wide, and no correction for multiplicity was applied. Similarly, the adjusted estimate for constructional praxis impairment was unstable because of the small number of events and should not be interpreted as evidence of harm. Overall, the most clinically coherent signal was the lower proportion of patients with impairment in more than one cognitive domain among RIperC-treated patients, suggesting a possible reduction in global cognitive burden rather than a robust effect on a single isolated domain.

This substudy has several limitations. First, the sample size was modest, and only 58 of 122 patients included in the primary REMOTE-CAT analysis completed the 90-day neuropsychological assessment. Patients included in the cognitive substudy had lower admission NIHSS scores and were more likely to achieve favorable functional outcome at 90 days than those not included, indicating selection toward patients with less severe strokes and better recovery. However, treatment allocation was well balanced between included and nonincluded patients, suggesting that this selection was related to baseline severity, recovery, and logistical factors rather than to the randomized treatment group. Second, the cognitive substudy was performed at a single participating center, limiting generalizability and introducing potential center-specific selection. Finally, the small number of impairment events in several domains limited the precision of adjusted estimates.

Despite these limitations, this substudy provides clinically relevant information. It shows that standardized cognitive assessment can be incorporated into a prehospital acute stroke neuroprotection trial and suggests that cognitive outcomes may reveal signals not fully captured by global disability scales. Although prehospital RIperC did not significantly improve 90-day cognitive outcomes in this exploratory analysis, the numerical trends observed for global cognitive burden and visual memory support further investigation. Future ischemic conditioning trials should consider incorporating prespecified cognitive endpoints, multicenter neuropsychological assessment, baseline or early poststroke cognitive measures, and longitudinal follow-up to better determine whether RIperC can influence poststroke cognitive recovery.

## Data Availability

The corresponding author will consider reasonable requests for access to the deidentified data reported in this article, subject to approval according to applicable ethical, legal, and data protection requirements.

## Acknowledgments

The authors thank the REMOTE-CAT Trial Investigators and all emergency medical services professionals and ambulance technicians involved in the trial. The REMOTE-CAT Trial Investigators include Cristina García-Vázquez, Yhovani Gallego, Maria Pilar Gil-Villar, Cristina González-Mingot, Alejandro Quílez, Emilio Ruiz-Fernández, Ignacio Saldaña-Inda, Jordi Sanahuja, Agustín Sancho-Saldaña, Eduardo San Pedro, Ana I. García-Díaz, Albert Freixa, Maria Viladrosa, Silvia Gros, Maria Muchada, Andrés Paipa, Carla Avellaneda-Gómez, Juan J. Mengual, Pere Cardona, Manuel Gómez-Choco, Jorge Pagola, and Sònia Abilleira.

## Contributors

FP and GM had full access to all data in the study and take responsibility for the integrity of the data and the accuracy of the data analysis. Concept and design: FP, GA, GM, XJF. Acquisition, analysis, or interpretation of data: FP, GA, CP. FP and CP wrote the first draft of the report. Critical revision of the manuscript for important intellectual content: FP, CP, GA, AR, XJF, TS, JRR, MV, AR, SS, CGV, RCS, GMC. Statistical analysis: RCS. Obtained funding: FP. Supervision: FP, GA. All authors had full access to all the data in the study and had final responsibility for the decision to submit for publication.

## Data Sharing Statement

The corresponding author will consider requests for access to the data reported in this article.

## Use of Artificial Intelligence

During preparation of this manuscript, ChatGPT, developed by OpenAI, was used solely to assist with grammar, style, and language refinement. This assistance did not affect the scientific content, data analysis, data interpretation, or study conclusions.

